# Single cell gene expression profiling of nasal ciliated cells reveals distinctive biological processes related to epigenetic mechanisms in patients with severe COVID-19

**DOI:** 10.1101/2021.12.08.21267478

**Authors:** Luis Diambra, Andres M Alonso, Silvia Sookoian, Carlos J Pirola

## Abstract

**Objective:** To explore the molecular processes associated with cellular regulatory programs in patients with COVID-19, including gene activation or repression mediated by epigenetic mechanisms. We hypothesized that a comprehensive gene expression profiling of nasopharyngeal epithelial cells might expand understanding of the pathogenic mechanisms of severe COVID-19.

**Methods:** We used single-cell RNA sequencing (scRNAseq) profiling of ciliated cells (*n* = 12725) from healthy controls (SARS-CoV-2 negative *n* =13) and patients with mild/moderate (*n* =13) and severe (*n* =14) COVID-19. ScRNAseq data at the patient level were used to perform gene set and pathway enrichment analyses. We prioritized candidate miRNA-target interactions and epigenetic mechanisms.

**Results:** Pathways linked to mitochondrial function, misfolded proteins, and membrane permeability were upregulated in patients with mild/moderate disease compared to healthy controls. Besides, we noted that compared to mild/moderate disease, cells derived from severe COVID-19 patients had downregulation of sub-networks associated with epigenetic mechanisms, including DNA and histone H3K4 methylation and chromatin remodeling regulation. We found 11-ranked miRNAs that may explain miRNA-dependent regulation of histone methylation, some of which share seed sequences with SARS-CoV-2 miRNAs.

**Conclusion:** Our results may provide novel insights into the epigenetic mechanisms mediating the clinical course of SARS-CoV-2 infection.

## INTRODUCTION

Cumulative and consistent evidence based on the records of patients with COVID-19 demonstrated that the disease course is characterized by a complex clinical spectrum, including asymptomatic infection-which is often not fully captured by cross-sectional epidemiological surveys-, mild symptoms, critical illness, and even the so-called long-COVID-19[1–4]. While progress has been made to understand the mechanism/s behind the evolution of the disease from mild to severe clinical outcomes, the molecular processes associated with the worst COVID-19 phenotypes, including severe pneumonia, acute respiratory distress syndrome, and even systemic organ failure, are not completely understood.

SARS-CoV-2 disseminates by exposure to infectious respiratory fluids, and before symptoms onset, the viral load in cells of nasopharyngeal epithelium is usually high [5]. Therefore, the functional role of the nasal epithelial cells not only as of the first physical barrier against viral entry but also by controlling the local immune response is of utmost importance in determining the disease course.

Single-cell RNA sequencing (scRNA-seq) profiling has allowed the identification of genes and molecular pathways associated with diverse human conditions, including pathways that define how a given cell can modify its phenotype after specific stimuli like a viral infection. Thus, scRNA-seq is a state-of-the-art approach for exploring changes in cellular regulatory programs, including activation or repression of genes that may explain the pathophysiology of human disease traits. For example, Ahn and coworkers recently demonstrated that SARS-CoV-2 is massively replicated in a distinct subset of human nasal epithelial cells[6]. Furthermore, by scRNA-seq and *in situ* mappings of SARS-CoV-2 entry–related host molecules, the investigators showed that multiciliated cells but not secretory or basal cells of human nasal respiratory epithelium are the primary target for SARS-CoV-2 replication [6].

Furthermore, Ziegler et al., by performing elegant scRNA-seq profiling on nasopharyngeal cells from healthy and COVID-19 participants, revealed key mechanisms that may define protective or detrimental responses to SARS-CoV-2 [7]. Specifically, the authors demonstrated impaired immune response and anti-viral immunity in cells of the nasal epithelium along 18 different clusters - from inflammatory macrophages to epithelial cell identities that underline severe COVID-19 [7]. Likewise, Ziegler and coworkers mapped the developmental transitions among nasal epithelial cells from healthy epithelium to the epithelium of patients with severe disease [7]. The authors also provided compelling evidence on the mechanisms of host-SARS-CoV-2 interaction in terms of phenotypic differences among cell subtypes and the anti-viral and interferon-mediated response curbing the intracellular levels of viral replication [7]. However, the molecular processes of gene transcriptional regulation in severe COVID-19, particularly those under the influence of miRNAs, have not been fully elucidated. Here, we leveraged scRNA-seq open data to expand the analysis of biological processes and gene regulation programs associated with severe COVID-19.

## METHODS

### Biological samples

Our study comprises data from samples of the nasopharyngeal epithelium of 50 individuals from the University of Mississippi Medical Center recruited between April and September 2020 [7]. The sample included 15 healthy subjects with negative SARS-CoV-2 PCR tests, denoted by healthy control (HC), and 35 individuals diagnosed with COVID-19 and of whom nasopharyngeal swabs were collected within the first three days following admission to the hospital. Based on the World Health Organization (WHO) guidelines for COVID-19 severity stratification, these 35 patients with positive SARS-CoV-2 PCR tests were divided into two subgroups: 14 individuals with mild/moderate COVID-19 (WHO scores equal to 1-5, COVID-19 M), and 21 individuals with severe COVID-19 (WHO scores equal to 6-8, COVID-19 S).

### Single-cell RNA-seq data analysis

We used the single-cell RNA-seq data from nasopharyngeal swabs publicly available for download and visualization via the Single Cell Portal: https://singlecell.broadinstitute.org/, under the identification SCP1289. Specifically, we focused our analyses on cells of the respiratory epithelium located in the nasal mucosa, and the transcriptomic profile of 32871 genes in 32588 cells was obtained. Owing to the typical dropout of single cells, many of these genes have zero counts in many cells. Analyzing these data is a real challenge. In our study, we considered only those genes with a non-null record in at least 650 cells (2 % of all cells). Thus, with this restriction, the present study included 11,870 filtered genes. The original study by Ziegler et al. [7] classified cells into 18 cell types (basal cells, B cells, ciliated cells, dendritic cells, deuterosomal cells, developing ciliated cells, developing secretory and goblet cells, enteroendocrine cells, erythroblasts, goblet cells, ionocytes, macrophages, mast cells, mitotic basal cells, plasmacytoid DCs, secretory cells, squamous cells, T cells). In the present study, we only considered the groups of ciliated cells, which include “ciliated cells,” “deuterosomal cells,” and “developing ciliated cells,” representing a set of 12725 cells. Unlike previous studies where differentially expressed genes (DEG) are determined considering “each cell” as an independent sample or replicate, we considered “each individual or patient” as an independent replicate. Thus, we calculated the mean expression level of each gene in each individual by averaging the normalized counts of all cells of the specific ciliated cell type belonging to that individual. In this manner, we avoided inflating statistical differences as the inclusion of the whole single-cell data taking individual cells as biological replicates might result in an overestimation of significant DEG. The number of cells of a given type varies from patient to patient; some are rare in some individuals. In this study, only those patients with more than 45 ciliary cells were considered.

### Pathway enrichment analysis and data visualization

The scRNA-seq data derived from the different groups of patients were contrasted in three different ways: 1) functional analysis of biological processes (BPs) comparing HC vs. COVID-19 M, and COVID-19 S vs. COVID-19 M using GSEA, as described below; 2) analysis of regulatory target gene sets associated with miRNAs contrasting COVID-19 S vs. COVID-19 M using SEA; and 3) gene-set enrichment tools by the ToppGene Suite available https://toppgene.cchmc.org/.

First, we analyzed the GO (gene ontology) terms for BPs associated with DEG using GSEA (Gene Set Enrichment Analysis software, https://www.gsea-msigdb.org) [8]). Briefly, GSEA is a computational method that determines whether a ranked gene list shows statistically significant, concordant differences between two phenotypes. Here, the rank of genes was established by the Signal2Noise metric, which is defined as the difference between the means expression in each phenotypic class divided by the sum of the deviations as implemented in the GSEA platform for dichotomic phenotypes. We performed the comparisons considering a subset of the C5 gene set (version 7.4, from MSigDB collection) that includes the BP ontology and a gene set size in the range of 10-500 genes. Briefly, GO sets are based on ontologies and do not necessarily comprise co-regulated genes. In all cases, we used 1000 permutations over genes to compute the statistical significance of enrichment scores. Next, we performed a cluster analysis of significant BPs by Enrichment Map Cytoscape App 3.3 [9] to obtain a BP network, where nodes represent BPs and edges represent gene overlap between gene sets associated with the connected nodes. Edges with a similarity score lower than 0.5 were not included in the BP network plot. Network clustering and annotation were performed by MCL (Markov Cluster algorithm), and Word Cloud (normalization factor 0.5) using the Cytoscape plug-in auto annotate [10].

Second, we used the GSEA algorithm to prioritize candidate miRNA-target interactions in patients with COVID-19 across the two primary clinical outcomes of interest, including patients with severe vs. patients with mild/moderate disease. To this end, we used the subset of the C3 gene sets (version 7.4, from MSigDB collection) that includes miRNA-targets genes as cataloged by the miRDB v6.0 algorithm [11]. We considered only gene sets with a size in the range of 300-1000 genes.

Finally, we performed gene list functional enrichment of a selected input list (see the results section for further details) to predict gene-drug interactions and co-expression analysis that was compiled from various data sources and used to build training set gene profile by the ToppGene/ToppCluster resource (https://toppgene.cchmc.org/) [12,13]. The same platform was used to interpret BPs, molecular functions, family genes, and human traits associated with particular subsets of genes or prioritize putative viral miRNA targets as described below.

## RESULTS

We noted that the number of cells of a given type varied considerably from patient to patient, and some cell types are rare in some individuals. Therefore, in this study, we only considered those patients that presented more than 45 ciliary cells. Consequently, we included in our analysis 13 healthy controls that were SARS-CoV-2 negative by PCR, 13 patients with mild/moderate COVID-19, and 14 patients with severe COVID-19.

### Gene set enrichment analysis associated with COVID-19 across all levels of the disease severity identifies distinctive biological processes

As explained earlier, we performed gene set enrichment analysis (GSEA) of BPs associated with SARS-CoV-2 infection in cells of the respiratory epithelium located in the nasal mucosa. Hence, we analyzed a total number of 12725 ciliated cells. We first compared BPs in infected patients presenting mild/moderated COVID-19 versus control (healthy) subjects. Besides the GO terms associated with innate immune response and TNF-mediate signaling, genome replication and viral release, viral budding and symbiont-host interaction, granulocyte migration, and lymphocyte homeostasis that were already described by Ziegler et al. [7], we found some other BPs and/or sub-networks that deserve further analysis (**Figure 1**). For example, we found that sub-networks associated with mitochondrial membrane permeabilization, protein targeting to mitochondria, and lysosome and protein targeting to the membrane were significantly enriched in patients with mild/moderate COVID-19. In addition, sub-networks related to protein folding and misfolding are highlighted. This last observation could be related to a previous characteristic described for SARS-CoV2 codon composition of highly expressed genes, which could influence the tRNAs pools of the infected cells with relevant implications in host protein folding and expression [14]. The entire list of BPs, including the GO terms and related genes, is shown in **Supplementary Table 1**.

**Figure 1:**
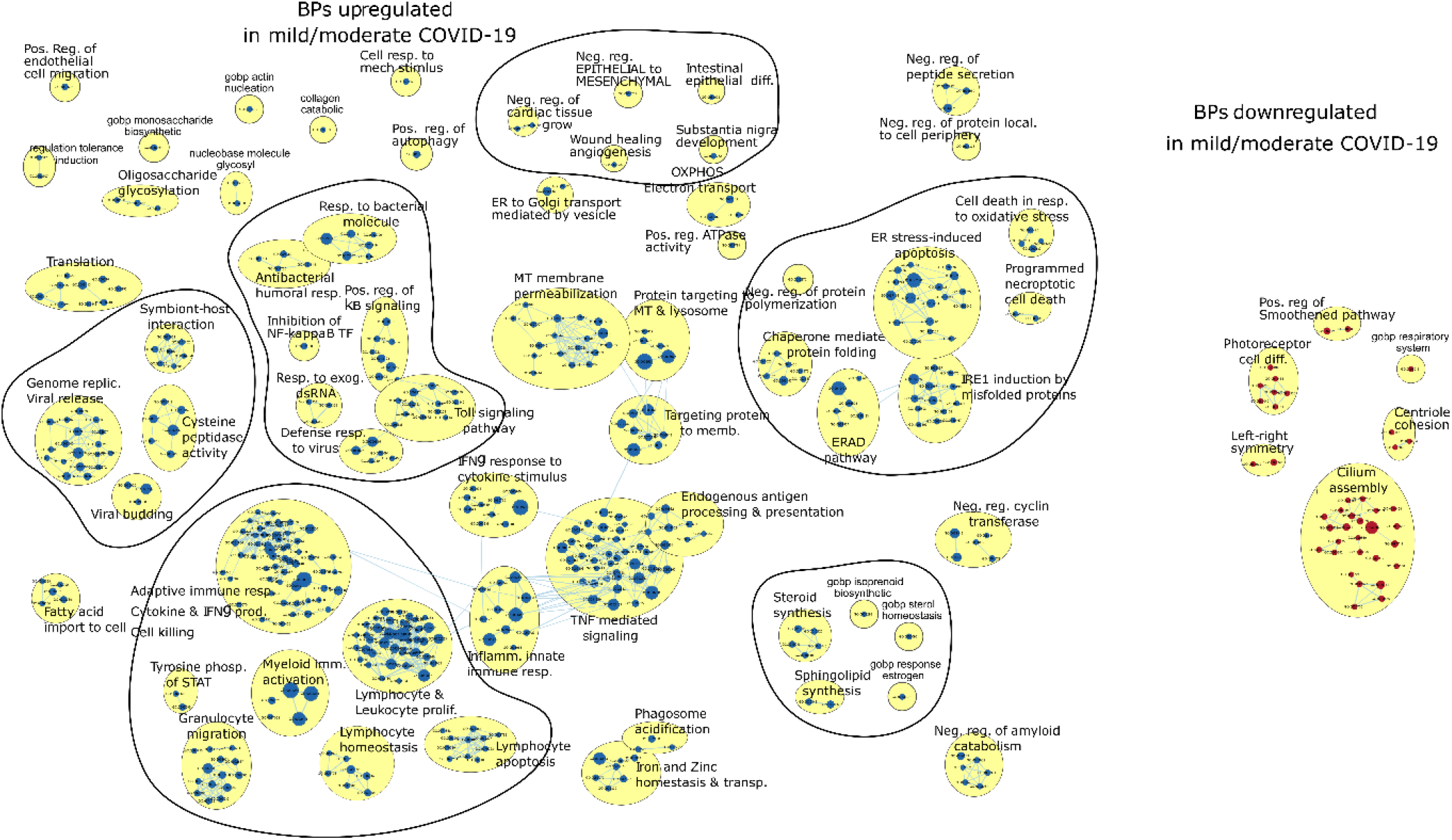
Gene set enrichment analysis derived from ciliated cells in mild/moderate COVID-19 patients vs. healthy controls. The Figure illustrates a network of GO (Gene Ontology) terms corresponding to biological processes (BPs) significantly enriched in genes upregulated (blue nodes) and downregulated (red nodes) in mild-moderate COVID-19 (COVID-M) patients with respect to healthy control (HC) individuals (Bonferroni adjusted p-value, FDR 0.05). The size of the node is proportional to the number of genes associated with that BP. The edges represent gene overlap between gene sets related to different GO terms. Connected nodes are organized in clusters of interconnected BPs obtained by the MCL algorithm, which considers similarity among gene sets to assign the edges, with a similarity score threshold of 0.5.

We next asked whether the differential pattern of BPs observed in ciliated cells may distinguish mild and moderate cases from severe COVID-19. The most relevant sub-networks identified by GSEA are shown in **Figure 2**. Compared to mild/moderate COVID-19 infection, patients with severe disease showed overrepresentation of sub-networks linked to fatty acid metabolism and eicosanoid synthesis, acute inflammatory processes, keratinocyte differentiation and humoral immune response, which were all described by Ziegler et al.[7]. However, we noted that cells derived from severe COVID-19 patients had some remarkable underrepresented sub-networks linked to epigenetic mechanisms, including histone H3K4 methylation and regulation of histone methylation (**Figure 2**). In addition, GO terms associated with ciliogenesis (GO:0003341, GO:0035082, GO:1905515, and GO:0001578) were significantly downregulated in severe COVID-19 (**Figure 2**). The entire list of BPs, including the GO terms and related genes, is shown in **Supplementary Table 2**.

**Figure 2:**
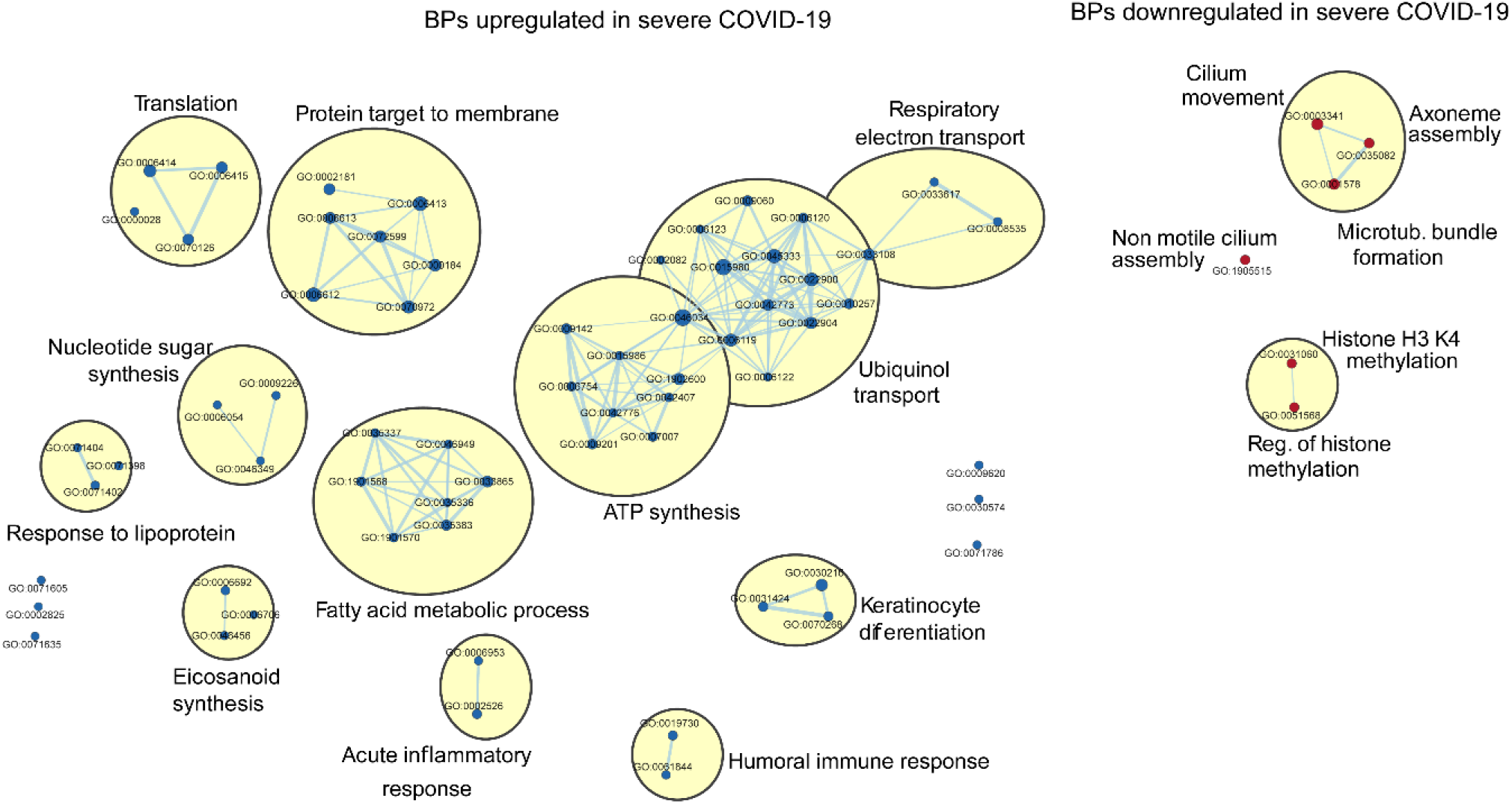
Gene set enrichment analysis derived from ciliated cells in severe vs. mild / moderateCOVID-19 patients. The Figure illustrates enrichment analysis of biological processes (BPs) upregulated (blue nodes) and downregulated (red nodes) in patients with severe COVID-19 vs. patients with mild/moderate disease(Bonferroni adjusted p-value, FDR 0.05). BPs were clustered by using the MCL algorithm with a similarity score threshold of 0.5.

### Gene expression profile of cells derived from the nasal mucosa in patients with COVID-19 across diverse stages of disease severity: sub-networks analysis focused on organelle function

To gain further insight into the gene lists generated by the GSEA analysis, we focus on specific pathways connected with impaired organelle function. Specifically, we selected the pathways of mitochondrial membrane permeabilization, protein targeting to mitochondria, and lysosome and protein targeting to the membrane, in which transcript levels of most upregulated genes in COVID-19 M patients with respect to control subjects are in the percentile 0.10. Then, we compared the expression profile of these genes within each experimental group, including HC, COVID-19 M, and COVID-19 S; transcript levels in each group of subjects were referred to the media of all samples. A heat map of the results appears in **Figure 3**. By including the COVID-19 S group, the analysis shows significant downregulation of certain genes in cells derived from COVID-19 patients with severe disease. For instance, the expression of *LCK* (Lymphocyte Cell-Specific Protein-Tyrosine Kinase, of which the encoded protein is a key signaling molecule in the selection and maturation of developing T-cells), *TP63* (a member of the p53 family of transcription factors), *TRAM1* (Translocation Associated Membrane Protein 1), and *SLC40A1* (Solute Carrier Family 40 Member 1) were significantly decreased in severe COVID-19. Upon inspecting the pattern of expression in members of the ribosomal protein family (RPL and RPS) that encode structural constituents of ribosomes, we found that cells of patients with COVID-19 had significant upregulation regardless of the disease severity. Interestingly, these ribosomal protein members are associated with other viral diseases, particularly influenza [15].

**Figure 3.**
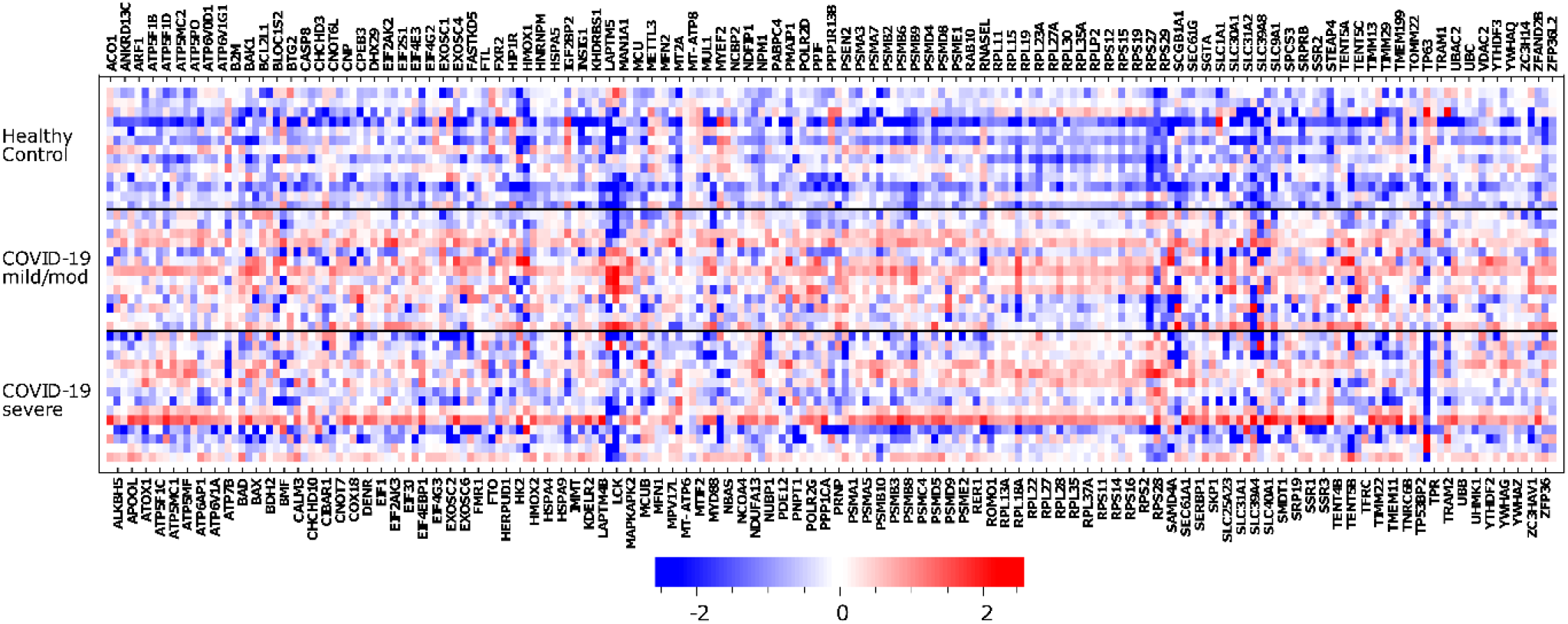
Heat map of expression genes belonging to the pathways associated with mitochondrial membrane permeabilization, protein targeting to mitochondria, and lysosome and protein targeting the membrane. Blue squares correspond to downregulated genes and red squares to upregulated genes. We selected for the analysis only the subset of genes most upregulated (the percentile 0.1 of the ranked genes)in severe vs. mild/moderate COVID-19 patients. The color scale stands for the RNA abundance relative to the media of transcript levels in all subjects and then log2 transformed.

### The regulatory network of miRNAs associated with severe COVID-19

We next sought to explore the putative regulatory mechanisms associated with more severe illness. Specifically, we asked whether miRNA-mediated deregulation of gene expression may explain severe COVID-19. Hence, using the ranked list of genes obtained by GSEA from the comparison of COVID-19 S vs. COVID-19 M as input, we detected 29 miRNAs potentially affecting the disease severity.

Some of these miRNAs share the same so-called seed region, which ultimately defined the mRNA target, and in that case, have the same gene targets. Therefore, we curated the list of miRNAs by an in-house script to avoid redundancy in the subsequent analyses. The correlations among the targets of the 29 miRNAs mentioned above are depicted in **Supplementary Figure 1**. Upon inspection of redundancy, 11 ranked miRNAs (miR-16-5p, miR-130-3p, miR548h-3p, miR-1283, miR548an, miR-19a-3p, miR144-3p, miR9983, miR101-3p, miR181a-5p, and miR8485) were used in the search for their targets among the whole set of putative miRNA target genes of the human genome (hsa-miRNA targets as cataloged by the miRDB v6.0 algorithm). As a result, we found a total of 3076 target genes. The complete list of target genes associated with the 11 miRNAs is shown in **Supplementary Table 3**.

In parallel, we selected the 10 % top-ranked genes downregulated in COVID-19 S vs. COVID-19 M according to the ranking based on the Signal2Noise metric, which yielded 1167 genes (**Figure 4a**). From the intersection of the 11 miRNAs-targeted genes (3076) and the above described 1167 downregulated genes in severe COVID-19, we found 336 deregulated transcripts that are also targets of the 11 miRNAs (**Figure 4b**). The expression pattern of these 336 genes within each group of patients relative to the media of all groups (healthy controls, COVID-19 M, and COVID-19 S) is shown in **Figure 4c**. A comprehensive network of the BPs represented by these downregulated genes in patients with severe COVID-19 is shown in **Figure 5**.

**Figure 4.**
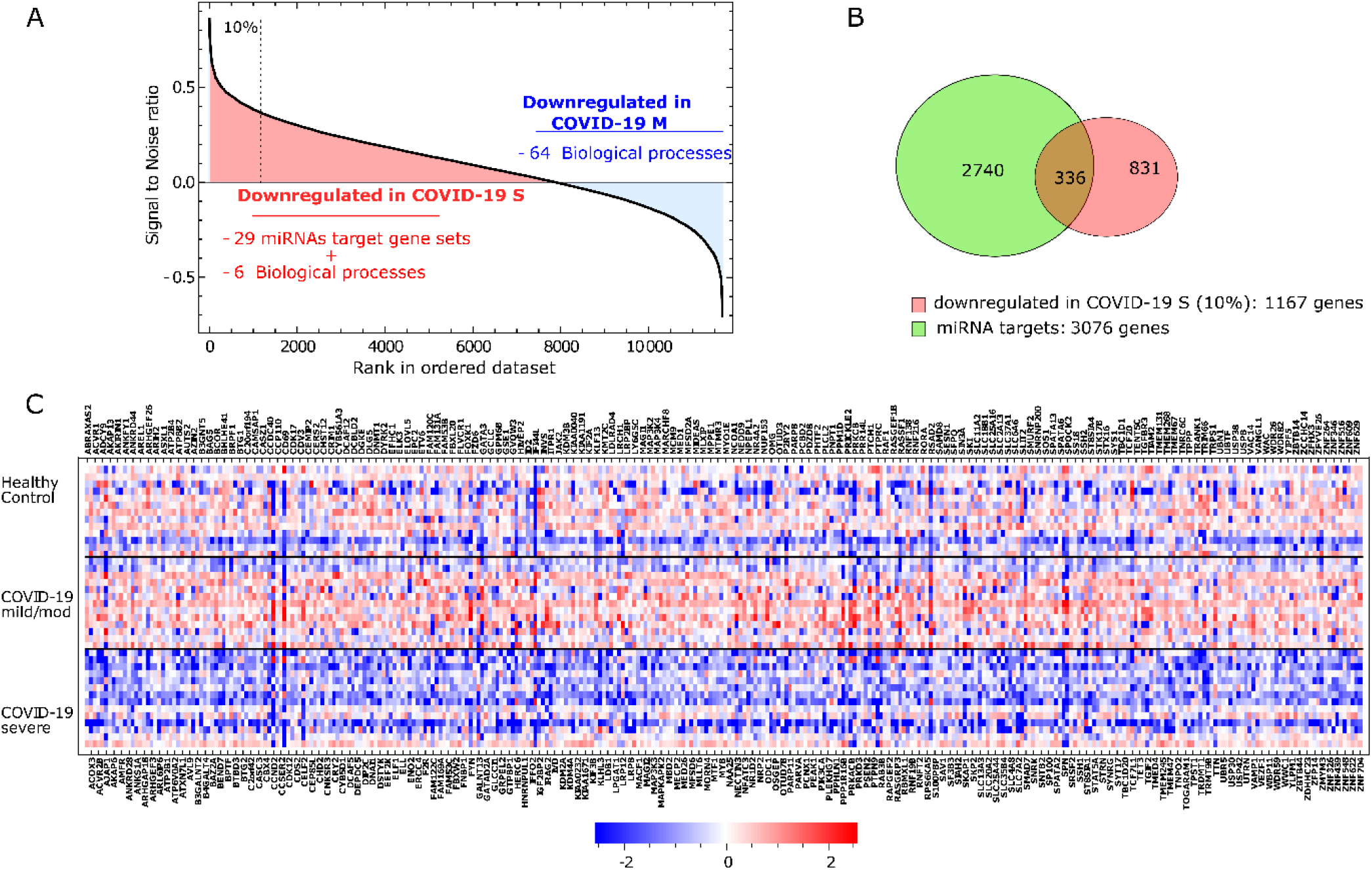
The role of epigenetic mechanisms in COVID-19: a regulatory network of miRNAs-target genes. A. Rank of genes according to the Signal2Noise metric as implemented in the GSEA platform (COVID-19 S vs. COVID-19 M) using the media of the normalized counts of the whole cell population in each individual. The top 10% of downregulated genes (rank over the dashed line) was used in the subsequent analysis showed in panels B and C. B. Venn Diagram showing the overlap between the 3076 gene targets of the 11 miRNAs associated with the target sets enriched in COVID-19 S vs. COVID-19 M and the 1167 top-ranked genes downregulated in COVID-19 S (as explained in panel A). The intersection shows the 336 genes used in panel C. C. A heat map of the 336 genes that result from the intersection mentioned in panel B. Squares in blue and red represent genes with lower and higher RNA abundance relative to the media of transcript level of all subjects and then log2 transformed, respectively. COVID-19 S: severe disease; COVID-19 M: mild/moderate disease.

**Figure 5.**
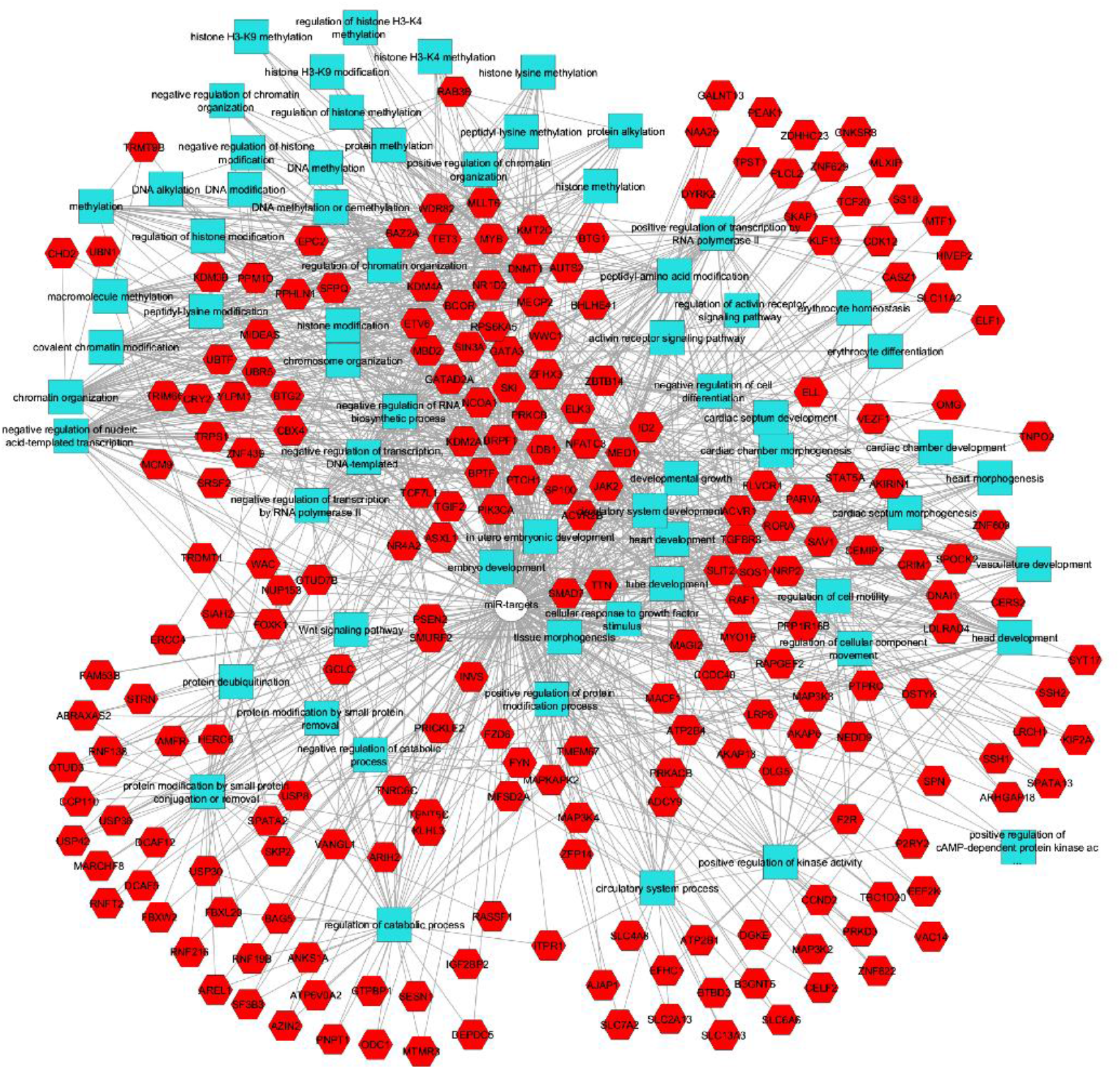
Network of the 336 genes downregulated in COVID-S and their associated GO terms for Biological processes (BP) The network was constructed using the ToppGene/ToppCluster resource (https://toppgene.cchmc.org/) with an FDR of 0.05. Hexagons in red and squares in light blue stand for genes and BPs, respectively, which were distributed using the edge-weighted Spring embedded layout and slightly modified for readability using Cytoscape v3.4.0. In general, the upper left region shows BP associated with DNA and histone modifications, regulation of transcription, and chromatin or chromosome remodeling. In contrast, the right middle part shows BPs associated with organ morphogenesis, particularly cardiac and vascular. The lower left region shows metabolic processes, primarily catabolic.

We then focused our analysis on those miRNA-target pairs involved in epigenetic mechanisms, including BPs linked to H3K4 methylation and regulation of histone methylation. In fact, these 11 miRNAs together regulate most of the histone methylation pathway genes that were downregulated in severe COVID-19 (*MYB*, BCOR, *DNMT1, KMT2C, TET3, KMT2A, KMT2C, WDR82, GATA3, AUTS2, MECP2, and MLLT6*) (Supplementary Figure 2).

### Gene list enrichment analysis based on functional annotations: insights into drug-target and co-expression functional enrichments

We finally sought to explore potential druggable targets in the list of differentially expressed miRNA-target genes associated with epigenetic mechanisms.

The transcripts in the input list (*MYB*, BCOR, *DNMT1, KMT2C, TET3, KMT2A, KMT2C, WDR82, GATA3, AUTS2, MECP2, and MLLT6*) were then screened for gene-drug interactions using the ToppGene Suite resource.

We found several compounds as putative ligands of the genes enriched in the input list; however, some specific drugs were ranked among the top ones. These drugs included diethylstilbestrol (FDR=2.904E-2), trichostatin A (FDR= 1.902E-2), hydralazine hydrochloride (FDR= 1.902E-2), and chlorambucil (FDR= 1.902E-2). In addition, we found that a large proportion of the predicted tissue co-expression of the target genes was mainly located in cells of the immune system, including alpha-beta T cells and gamma-delta T cells of the pancreas, thymus, lymph node, spleen, and CD4 positive of the spleen (FDR 1.209E-2).

## DISCUSSION

In this study, we identified distinctive biological processes related to epigenetic mechanisms in patients with severe COVID-19. Specifically, we analyzed scRNA-seq profiles from nasopharyngeal ciliated cells of patients with COVID-19 and performed pathway enrichment analysis. Our research was based on two distinctive strategies, including the analysis of scRNA-seq data at an individual-patient level rather than at bulk cell-level and prioritization of molecular mechanisms associated with severe COVID-19.

We found that patients with severe COVID-19 presented deregulation of gene expression in pathways linked to mitochondrial function and membrane permeability. The expression of certain genes, including *LCK-*a tyrosine kinase involved in T-cells maturation and expansion, the tumor encoding protein *TP63* that plays a role in the regulation of epithelial morphogenesis, *TRAM1* that encodes for a multi-pass membrane protein that is part of the mammalian endoplasmic reticulum, and *SLC40A1* that mediates iron ion transmembrane transporter activity*-*among other transcripts, were consistently decreased across ciliated cells from all severe COVID-19 patients. Collectively, these findings reinforce the importance of ciliated cells differentiation, maturation, and immune response in these specific cells of the nasopharyngeal epithelium as highlighted by Ziegler et al. [7]. In addition to the relevance of mitochondrial dynamics, impairment of mitochondrial membrane potential, and cell death in the process of any viral infection, including SARS-CoV-2 infection, one may speculate that some other molecular mechanisms are relevant to determine the worst disease outcome. For instance, Singh et al. reported that SARS-CoV-2 interferes with mitochondrial function to evade host cell immunity and facilitate virus replication and COVID-19 development [16]. Likewise, disturbances in iron transport may affect mitochondrial function and induce oxidative stress. Earlier reports demonstrated that hyperferritinemia is a predictor of increased mortality of the disease [17]. Besides, ferroptosis, an iron-related cell death program, may be involved in COVID-19-induced multiple organ failure [18]. Furthermore, our results highlight the importance of ciliated cells in tissue homeostasis. Nasopharyngeal ciliated cells of patients with severe COVID-19 presented significant downregulation of pathways associated with cilium movement, microtubule bundle formation, and axoneme assembly. This finding has implications for understanding impaired physiological processes of the nasal mucosa, for instance, olfaction-related disorders and systemic functioning deregulation of ciliated cells across the body, including primary cilia in testicular cells. In fact, recent clinical evidence suggests that the spermatogenic function of the testis is impaired in patients with COVID-19 [19]. It is then plausible to speculate that the dysfunction of ciliated cells in severe COVID-19 underlies pleiotropic effects in many different tissues and organs of the body. Here, we found that in this phenomenon, the dynein protein family may play a crucial role, a finding already reported by others [20]. Therefore, the associated signaling pathways would resemble the syndromic disorders known as ciliopathies.

We also noted that cells derived from severe COVID-19 patients had downregulation of sub-networks linked to epigenetic mechanisms, including histone H3K4 methylation and regulation of histone methylation. As shown in **Figure 5**, genes in the pathways of histone modifications, histone lysine methylation, DNA methylation or demethylation, DNA modification, chromatin remodeling, chromosome organization, negative regulation of RNA biosynthetic process, negative regulation of the cellular biosynthetic process, and even cardiac morphogenesis - among many other pathways, are all dramatically downregulated in cells derived from severe COVID-19 patients. Some genes in this network belong to the Methyl-CpG binding domain-containing NuRD complex, Zinc fingers C2HC-type PHD finger proteins, Lysine acetyltransferases, and Zinc fingers CXXC-type gene families. Collectively, these findings might explain the differences in developmental and maturation trajectories of ciliated cells in severe COVID-19 patients reported by Ziegler and coworkers [7].

To expand the understanding of the potential molecular processes involved in the regulation of gene expression, we examined the enrichment of miRNAs in the list of genes deregulated in cells of patients with severe vs. mild/moderate COVID-19. We found some miRNA-gene-target pairs that might be relevant to the disease biology. For example, we highlight the miRNA-548, which has been shown to down-regulate host anti-viral response via direct targeting of IFN-λ1 [21]. Notably, miRNA-548 seems to participate in numerous signaling pathways, such as the Wnt signaling pathway, the MAPK and TGF-β signaling pathways, and regulation of the immune system in the transition from immune tolerance to immune activation in chronic hepatitis B [22]. Notably, predictions of putative small ligands of genes enriched in the input list of predicted miRNAs showed some interesting gene-drug interactions that might explain differences in clinical outcomes mediated by epigenetic mechanisms or help treat the disease. For example, a significant interaction between five of the target genes of the list of enriched miRNAs was predicted with diethylstilbestrol. It appears that estrogen treatment silences the inflammatory reactions and decreases virus titers leading to improved survival rate [23]. Previous experimental studies showed that 17β-estradiol protects females against influenza by recruiting neutrophils and increasing virus-specific CD8 T cell responses in the lungs [24]. Perets and colleagues showed that 17β-estradiol reduces influenza A virus replication in primary human nasal epithelial cells derived from females [25]. Some other gene-drug target predictions suggest that compounds used to treat hematological conditions, such as leukemia and lymphomas, or the antihypertensive agent hydralazine hydrochloride could be tested for drug repurposing in the treatment of severe COVID-19.

As a putative mechanistic explanation, we observed some overlap between the seeds of our DEG-associated miRNAs and those of the recently reported SARS-CoV-2-encoded viral miRNAs[26–28], as shown in **Supplementary Table 4**. Thus, these findings could indicate that SARS-CoV-2 has miRNAs-like sequences [28], and - like many other viruses-, hitchhikes the regulatory processes of the host cells by targeting epigenetic modulators. We are aware that our research may have limitations. First, the nature of retrieved metadata and the limited number of patients do not allow us to examine relevant demographic information of the infected patients. Therefore, we did not examine the correlation between the patients’ features, such as age and gender, with deregulated gene expression pathways. Second, the analyzed dataset contains scRNA-seq data from the nasopharyngeal swabs collected within the first three days following admission to the hospital [7], which does not allow any assessment of patient’s follow-up. Hence, further validation of our observations using experimental models either in cell lines or animal models is required. Together, our results may provide novel insights into the cellular and molecular mechanisms that modulate the clinical course of SARS-CoV-2 infection. Based on predicted biological processes and functional enrichment analysis, we detected some relevant mechanisms that may explain the pathogenesis of severe COVID-19. Among these molecular explanations, we found some clues on the importance of post-transcriptional regulation of gene expression by small-non coding RNAs and epigenetic mechanisms of gene transcription regulation, such as DNA and histone methylation and ultimately chromatin remodeling.

## Data Availability

All data produced in the present work are contained in the manuscript

## Abbreviations

BP: biological process
DEG: differentially expressed genes
GSEA: Gene set enrichment analysis
GO: gene ontology
miRNA: microRNA
scRNA-seq: single-cell RNA sequencing

## Conflict of interest statement

Nothing to disclosure

## Acknowledgment

This study was partially supported by grants PICT 2018-889 and PICT 2019-0528 (SS), PICT 2016-0135 and PICT2018-0620 (CJP), PICT 2018-03713 (LD) (Agencia Nacional de PromociónCientífica y Tecnológica, FONCyT), CONICET Proyectos UnidadesEjecutoras 2017, PUE 0055 (SS, CJP).

## SUPPLEMENTARY MATERIAL

**Supplementary Figure 1.**
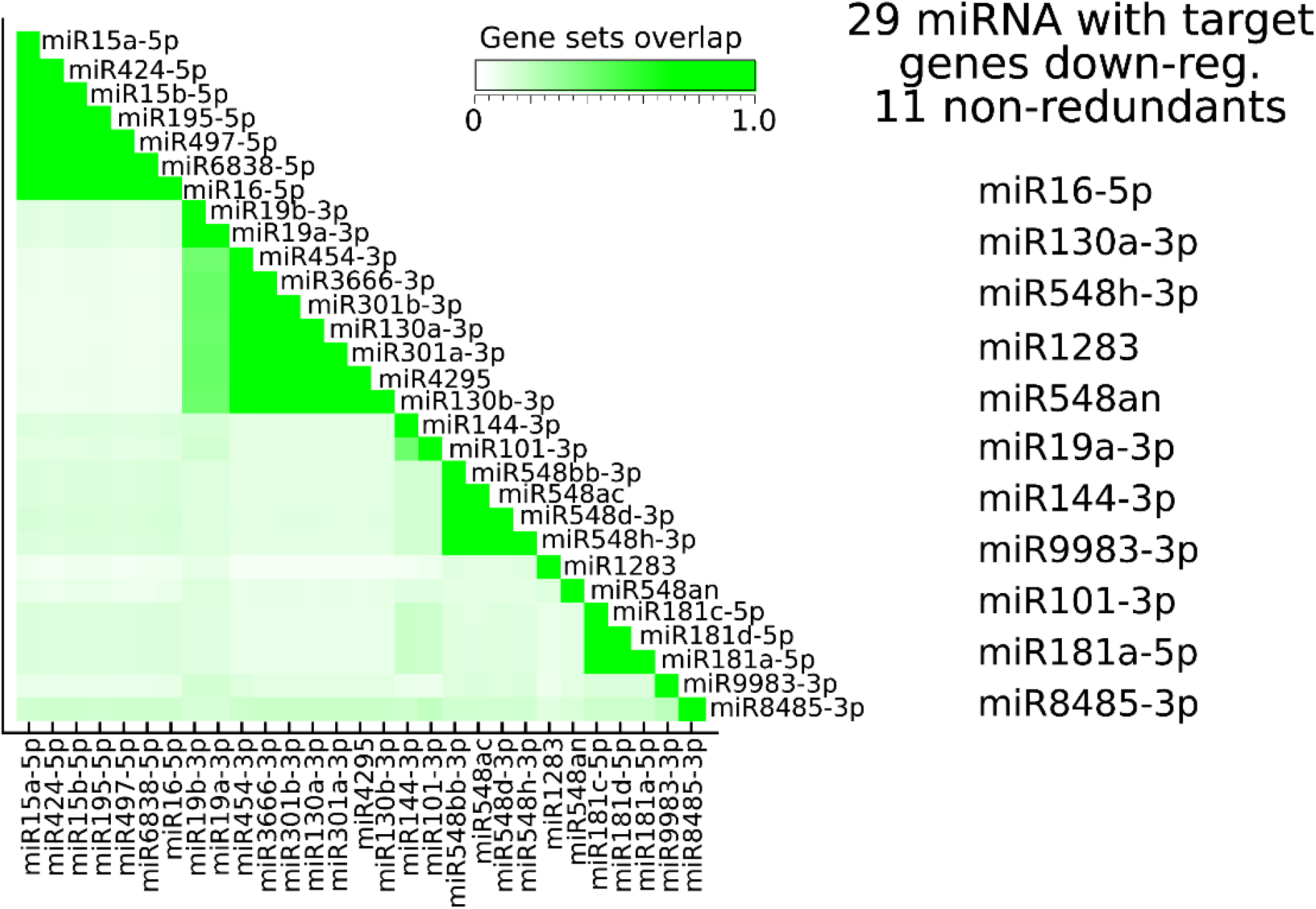
The regulatory network of miRNAs associated with severe COVID-19: correlation analysis. The comparative analysis of GSEA between cells derived from patients with mild and severe COVID-19 shows that DEG enriched in the group of mild patients (FDR <0.05) are the targets of 29 miRNAs. This Figure shows the fraction of target genes in common among the associated target genes of each of the 29 miRNAs. The miRNAs associated with each target gene sets show high redundancy among each other (green clusters). Therefore, there are only 11 target gene sets that have low redundancy among themselves. For the subsequent analyzes, we selected the corresponding 11 miRNAs listed to the right.

**Supplementary Figure 2.**
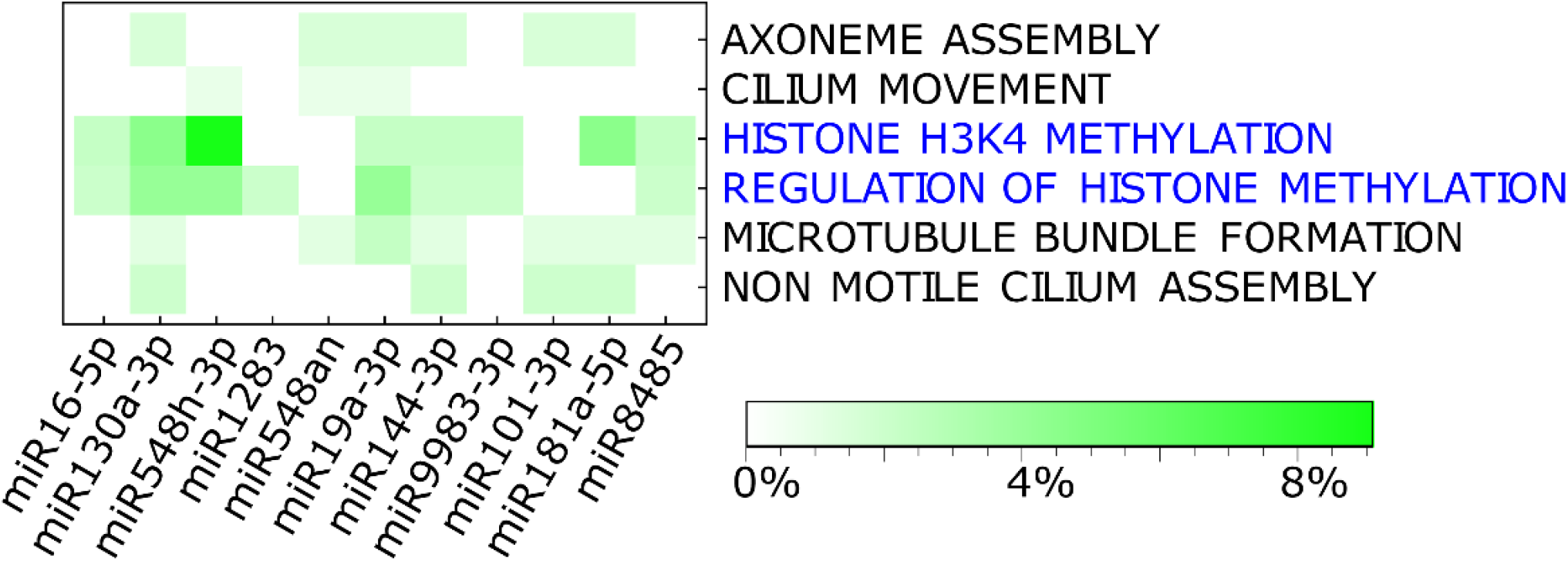
miRNA-target pairs involved in epigenetic mechanisms. The heat plot shows the percentage of overlap (green scale) between the target genes associated with each miRNA, with the genes involved in each downregulated BP in patients with severe COVID-19 shown in Figure 2 (FDR <0.05) and listed in Supplementary Table 2. Only those genes within the 10% most enriched genes in mild patients were taken into account. It is observed that most of the genes linked to histone methylation processes (highlighted in blue font) and that were downregulated in patients with severe COVID-19 are also target genes of miRNAs, which suggests that the presence of these miRNAs (or their viral homolog’s) deregulates histone methylation in these patients.

**Supplementary Table 1**

**Biological processes in mild/moderate COVID-19 vs. healthy controls**.

Biological processes down- (red rows) and upregulated (blue rows) in COVID-M with respect to HC as indicated by GSEA analysis (FDR<0.05), displayed in Figure. 1. The last column lists the genes associated with the GO terms. Enrichment score (ES) reflects the degree of overrepresentation of a gene set at the ranked list of genes. NES is the ES normalized across analyzed gene sets. FWER stands for family wise-error rate.

**Supplementary Table 2**

**Biological processes in severe vs. mild/moderate COVID-19 patients**

Biological processes down- (red rows) and upregulated (blue rows) in COVID-19 S with respect to COVID-19 M as indicated by GSEA analysis (FDR<0.05), displayed in Figure 2. The last column lists the genes associated with the GO terms. Enrichment score (ES) reflects the degree of overrepresentation of a gene set at the ranked list of genes. NES is the ES normalized across analyzed gene sets. FWER is the family wise-error rate.

**Supplementary Table 3: miRNAs with target genes downregulated in severe COVID-19** miRNAs with target genes downregulated in COVID-19 S with respect to COVID-19 M as indicated by GSEA (FDR<0.05) after redundancy elimination. The last column lists the target genes associated with the specific miRNA. Enrichment score (ES) reflects the degree of overrepresentation of a gene set at the ranked list of genes. NES is the ES normalized across analyzed gene sets.FDR stands for false discovery rate.

**Supplementary Table 4: Sequences of SARS-CoV-2-encoded vi-miRNAs compared to hsa-miRNAs targeting DEG in COVID-19 S vs. COVID-19 M patients**

Local alignment between the 11 human miRNAs determined by GSEA and vi-miRNA sequences reported in three previous studies [26–28]. Shared sequences with 6 and 7 bp are highlighted in bold font. In addition, we highlighted in bold red font over the hsa-miRNA a defined seed shared by part of a vi-miRNA.

## Notes

### Competing Interest Statement

The authors have declared no competing interest.

### Funding Statement

This study was partially funded by:
Agencia Nacional de Promocion Cientifica y Tecnologica (Argentina)

### Author Declarations

This study involves only openly available human data, reported in a previous paper, which can be obtained from: https://singlecell.broadinstitute.org/single_cell/study/SCP1289/impaired-local-intrinsic-immunity-to-sars-cov-2-infection-in-severe-covid-19

